# Relationship between fine particulate matter (PM_2.5_) ambient air pollution exposure to cardiorespiratory and muscular fitness in adults from MEDELLIN cohort study, 2022-2023

**DOI:** 10.64898/2025.12.17.25342533

**Authors:** Duván Alexis Gómez-Castro, Ana Liseth Herrera-Gómez, Carolina Betancur-Figueroa, Juan Gabriel Piñeros-Jiménez, Jaime Alberto Gallo-Villegas

**Author notes:** Corresponding author (JGPJ). Applied Medicine for Physical Activity and Sport, Faculty of Medicine, Universidad de Antioquia, Carrera 51D # 62- 29, Medellin, Colombia Medellin, Colombia (Duván Alexis Gómez-Castro, Ana Liseth Herrera-Gómez, Jaime Alberto Gallo-Villegas). Health and Environmental Group Research, National School of Public Health, Universidad de Antioquia, Calle 62 # 52- 59, Medellin, Colombia (Carolina Betancur-Figueroa, Juan Gabriel Piñeros-Jiménez). These authors contributed equally to this work.

## Abstract

**Background:** Air pollution caused by fine particles has been recognized as a significant environmental risk factor. Over the past two decades, there has been a substantial increase in evidence on the impact of air pollutants on mortality and morbidity in vulnerable groups, such as children under 5 years of age, people over 60 years of age, and people with comorbidities, primarily in low- and middle-income countries. However, most studies have focused on nosologically identified respiratory and cardiovascular events. Our objective was to evaluate the relationship between PM_2.5_ ambient air pollution exposure and -physical fitness (PF), an attribute that determines an individuaĺs physical performance and general health, in adults from Medellin, Colombia.

**Methods:** A cross-sectional nested cohort study was conducted to establish exposure to PM_2.5_ by each participant’s residence address. Physical fitness was assessed using the Dundee step test, sit-to-stand muscle power test, and grip strength. Multiple linear regression models adjusted for personal medical history were constructed to evaluate the study relationship.

**Results:** 320 participants were included, with an average age of 60 ± 8.7 years and an annual PM_2.5_ ambient air pollution of 18.9 ± 1.75 µg/m^3^. People with PM_2.5_ exposures above the 75th percentile showed greater use of the heart rate reserve (ß coefficient= 7.11; 95%CI 1.11-13.12) and better relative muscle power (ß coefficient= 0.50; 95%CI 0.23). -0.77) and grip strength (ß coefficient= 3.59; 95%CI 1.63-5.56).

**Conclusions:** This is the first study to explore the relationship between ambient PM_2.5_ ambient air pollution and cardiorespiratory and muscular fitness in a Latin American city. Our results indicate that people with greater exposure to PM_2.5_ have worse cardiorespiratory fitness (CRF) and better muscular fitness (MF).

**Key points:** Physical fitness determines an individual’s physical performance and general health status and includes indicators for establishing a cardiovascular baseline.

Prolonged exposure to air pollution negatively affects cardiovascular health. However, few studies have analyzed physical fitness indicators in relation to this environmental risk factor, and none have been conducted in the Latin American population.

It was established that people with higher exposure to PM_2.5_ ambient air pollution at their residence address had worse cardiorespiratory fitness.

## INTRODUCTION

Particulate matter (PM) has been identified as a global environmental public health problem [1]. It is a complex mixture of organic and inorganic components that are classified according to their aerodynamic size into coarse (diameter less than 10 microns, PM_10_), fine (diameter less than 2.5 microns, PM_2.5_), and ultrafine (diameter less than 0.1 microns, PM_0.1_) [2]. Fine particle matter reaches the distal airways and even crosses the alveolar-capillary barrier. According to the 2019 Global Burden of Disease study, PM_2.5_ is the fourth-highest risk factor for global mortality [3] and is attributed to 5.5 million premature deaths. It increases the risk of cardiovascular and respiratory issues, and causes problems with other systems [4], including physical fitness (PF) [5].

PF is an attribute that determines an individuaĺs physical performance and general health status, with the most influential factors being cardiorespiratory fitness (CRF) and muscular fitness (MF). CRF reflects the potential of the systemic response to supply oxygen demand [6] and is a determining factor in predicting cardiovascular risk [7]. An inverse relationship has been described between this capacity and cardiovascular, neurological, and oncological morbidity and mortality [8]. Having less than five metabolic equivalents (METs) is related to higher mortality from all causes and confirms the value of CRF as a protective factor in health [9]. MF corresponds to the neurological and musculoskeletal capacity to perform work, comprising neuromotor control, muscle strength, and endurance. It is directly associated with functional status and is a recognized protective factor against all-cause mortality [10].

CRF and MF are robust indicators of health that account for global physiological functioning. Long-term exposure to air pollution, particularly PM_2.5_, can negatively affect physical conditions [11]. No studies have previously evaluated this relationship in the Latin American population. For Colombian cities, air pollution is one of the biggest environmental problems [12], and there are reports of increased morbidity due to cardiovascular diseases associated with PM_2.5_ exposures [13]. This study aims to analyze the relationship between long-term exposure to PM_2.5_ and CRF and MF in adults, assuming that PM_2.5_ exposure levels are inversely related to these indicators.

## METHODS

### Ethics statement

This research was approved by the Research Ethics Committee of the National Faculty of Public Health of the University of Antioquia in session 165 of May 23, 2017 (number: 21030002 – 00223 – 2019). All procedures followed the ethical standards of the Declaration of Helsinki and its 2013 adaptation [14]. All participants agreed to participate voluntarily and signed an informed consent form that allowed them to follow the cohort and their clinical histories.

### Design and study area

This study is part of a research project called Exposure to Air Pollutants and Cardiovascular and Respiratory Health in Medellin (MEDELLIN cohort study), which evaluated the effect of long exposure to ambient particulate matter on health in a population-based cohort. A cross-sectional study nested in a cohort analyzed the relationship between PM_2.5_ and physical condition indicators in people over 44 years of age. This was carried out in Medellin, the second-largest city in Colombia. Medellin has a population of 2,376,337 inhabitants and is located in an inter-Andean valley in the northwest of the country. One-third of the city’s population is over 50 years old [15]. The city’s elevation is approximately 1400 m above sea level, with temperatures varying between 16 °C and 28 °C. In 2021, 90% of the Air Quality Monitoring Network stations reported annual PM_2.5_ concentrations below 25 µg/m^3^, the maximum value allowed by the Colombian standard [16].

### Population

The participants were inhabitants of two sub-city units as defined by Gouveia et al. 2021 [17], which were chosen due to the fact that the geographic estimates of ambient PM_2.5_ concentrations were of a higher certainty in the LUR developed in the reference study [18]. Participants were selected through simple random sampling proportional to the population size from each zone, and lived in households within a 600-meter radius of the city’s Air Quality Monitoring Network (AQMN) monitoring stations (https://siata.gov.co/siata_nuevo/) [19].

Participants were included if they were over 44 years of age, had resided in the study areas for five years or more, had not been diagnosed with a serious or decompensated illness at the time of recruitment, and understood and agreed with the conditions of participation. The recruitment period for this study was between 01/03/2022 and 31/05/2023.

### Air pollution exposure

Annual concentrations of PM_2.5_ were estimated using the results obtained from the land use regression (LUR) models built for 2022 using the methodology described in the ESCAPE study [18]. In summary, the data from the AQMN daily averages obtained between January 1 and December 31 were used to calculate the annual averages for each monitoring site. Meteorological, land use, and traffic variables were obtained from the city’s geographic information systems and were evaluated as predictors of spatial variation in annual PM_2.5_ concentrations. Regression models were built using a supervised stepwise approach for circular buffers with different radii (100, 150, 300, and 500 m) through spatial intersections of the explanatory variables. The model was selected for its statistical significance and compliance with the specification criteria of the ordinary least squares method.

Annual ambient PM_2.5_ concentrations were assigned to each residential address using the “Extract Multiple Values to Points” spatial statistics tool in ArcGIS version 10.7 (Environmental Systems Research Institute, Redlands, CA). Exposure was classified according to percentiles (P): lowest exposure <P25, intermediate exposure between P25-P75, and highest exposure ≥P75.

### Clinical survey

Demographic, clinical, and habit data were collected from each participant. A physical examination (vital signs, anthropometric variables, semiological evaluation, and functional tests) was performed, and the Charlson index was calculated to assess survival according to the number and severity of comorbidities [20]. Self-reported physical activity was rated according to World Health Organization recommendations [21].

### Cardiorespiratory Fitness assessment

The Dundee protocol step test was performed after visual and auditory demonstration at a height of 17.5 cm and a rhythm of 23 cycles per minute for 2 minutes [22]. The heart rate at rest (HR_r_), at the end of the test (HR_et_), and after the first minute of recovery were recorded with a sensor band (POLAR FT1, from the company Polar Electro Oy, Kemple, Finland). The maximum heart rate in subjects with or without beta-blockade treatment was calculated according to their respective formulas [23] and the percentage of heart rate reserve (%HR_r_) using the following formula: %HR_r_= (HR_et_-HR_r_/ HR_r_) x 100. The %HR_r_ directly correlates with the oxygen consumption reserve and estimates CRF [24]. Participants were classified as: “best CRF” participants with a %HR_r_ below the sample median and the “worst CRF” participants with a %HR_r_ above the median or those unable to perform or complete the test.

### Muscular Fitness assessment

A 30-second sit-to-stand test was performed on a standardized chair (0.45 m high), with instructions for performing repetitions as quickly as possible with arms crossed over the chest to assess strength and endurance. The number of repetitions was used to calculate relative muscle power according to weight. Participants were classified as those with “adequate power,” with a value above the median, and “inadequate,” with a value below, considering the reference values for the Colombian population according to age and sex [25].

The isometric grip strength of both hands (grip strength) was measured using a digital dynamometer (brand: Gripx, model G-EH101, United States). With the participant sitting, with their elbow static at 90°, maximum grip force was applied to the dynamometer at the second joint of the fingers below the grip handle. Starting with the non-dominant hand, the maximum value obtained was recorded, and the average for both hands was calculated and categorized as “adequate” or “inadequate”. The reference for grip strength was the 25th percentile for the South American population according to sex, age, and body mass index [26].

### Statistical analysis

Univariate analysis was performed according to the normality analysis of the data distribution using the Shapiro-Wilk test. Demographic, anthropometric, and clinical variables were described using the mean ± standard deviation (SD) or the median and interquartile range (IQR). Categorical variables were described with their frequency and their respective percentages. Differences in %HR_r_, relative muscle power, and grip strength according to covariates such as sex and smoking habits (Student’s t-test), level of physical activity, and socioeconomic status (ANOVA) were established. The correlation of the outcomes with age, body mass index (BMI), and comorbidity index (Spearman correlation) was analyzed.

Multiple linear regression models were built to assess the relationship between long-term exposure to PM_2.5_ and the outcomes. The Hosmer-Lemeshow criterion and biological plausibility were used to select variables to be incorporated into the models. Three models were constructed: Model 1 included age, BMI, and sex; Model 2 consisted of the variables from Model 1 plus the comorbidity index and physical activity level; and Model 3 consisted of the variables from Model 2 plus socioeconomic status and smoking habits. The best model was selected using the Akaike information criterion and the deviance of the models. The collinearity of the variables included in the model was assessed using tolerance and the variance inflation factor. The Shapiro-Wilk test, the Breusch-Pagan test, and the Durbin-Watson statistic were used to analyze the normality, homoscedasticity, and autocorrelation, respectively, of the residuals. Binomial logistic regression models were performed, and the outcomes of interest were considered dichotomous variables (adequate and inadequate) for sensitivity analysis. These analyses were performed using the Stata 18 software from StataCorp LLC.

## RESULTS

### Participants

By May 2023, 1,997 people had been enrolled in the cohort, of whom 954 met the participation criteria. However, only 320 people agreed to the clinical evaluation (Figure 1). The participants’ mean age was 60 ± 8.7 years; 71.2% (n= 228) were women, and 22.5% (n= 72) were employed. The mean BMI was 28.3 ± 5.1 kg/m^2^, with 33.4% (n= 107) of participants diagnosed with obesity and 39.7% (n= 127) with high blood pressure. Regarding healthy lifestyle habits, 11.4% (n=43) smoked, and 61.9% (n=198) did not perform the amount of physical activity recommended for good health in the general population. A more detailed description of the participants’ characteristics is presented in Table 1.

**Figure 1:**
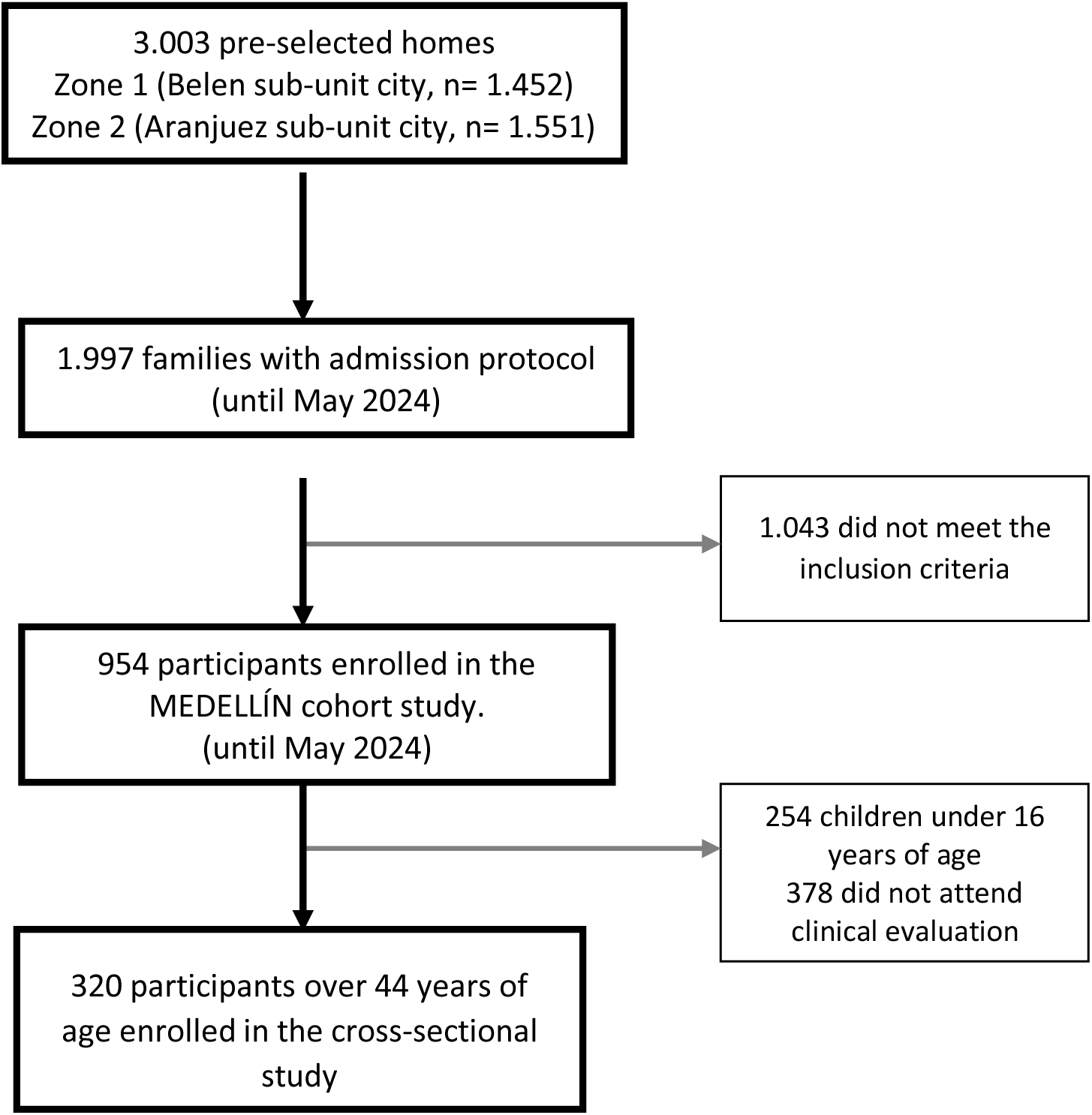
Participant selection process in the cross-sectional nested in the MEDELLIN cohort study.

**Table 1.**
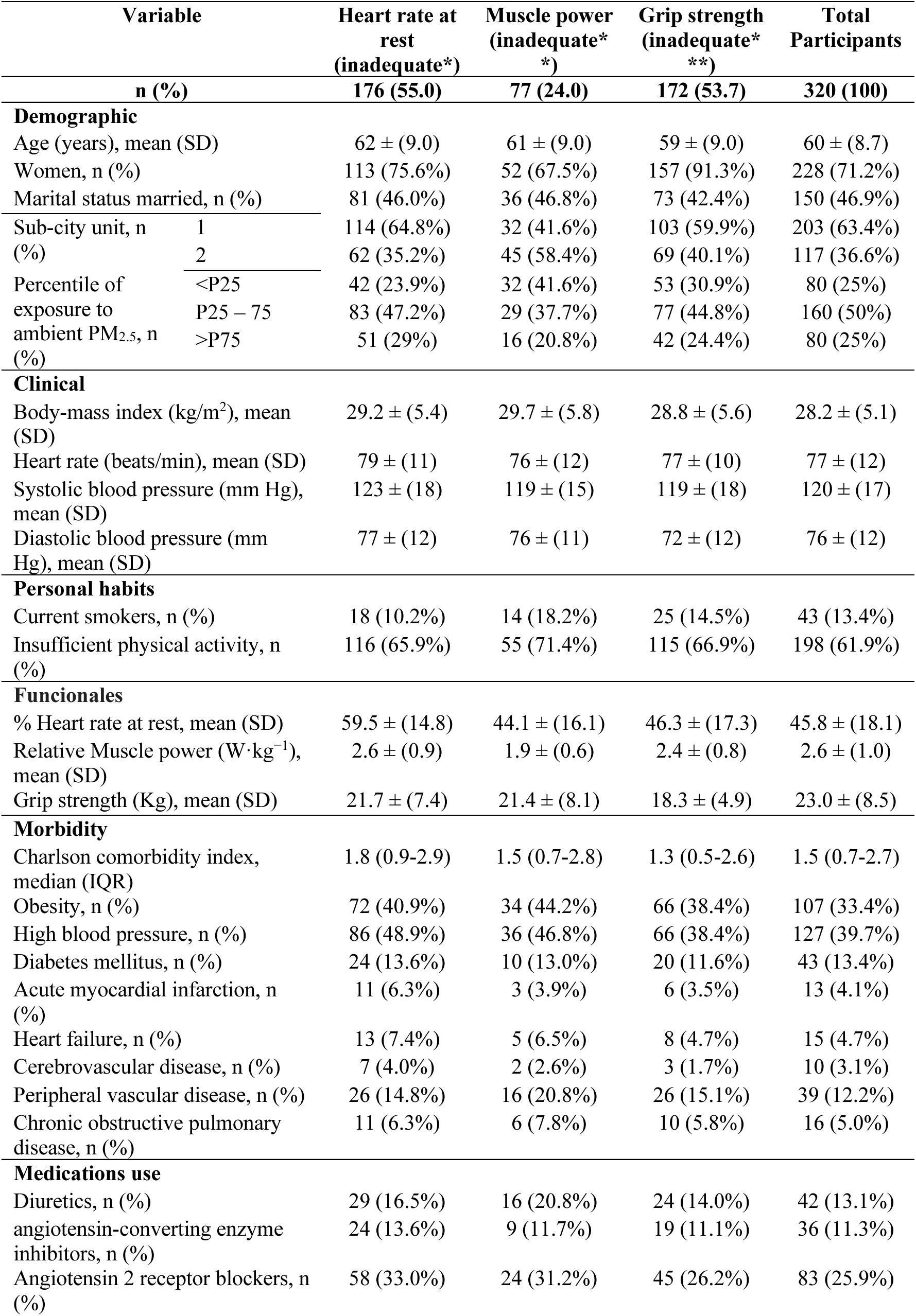
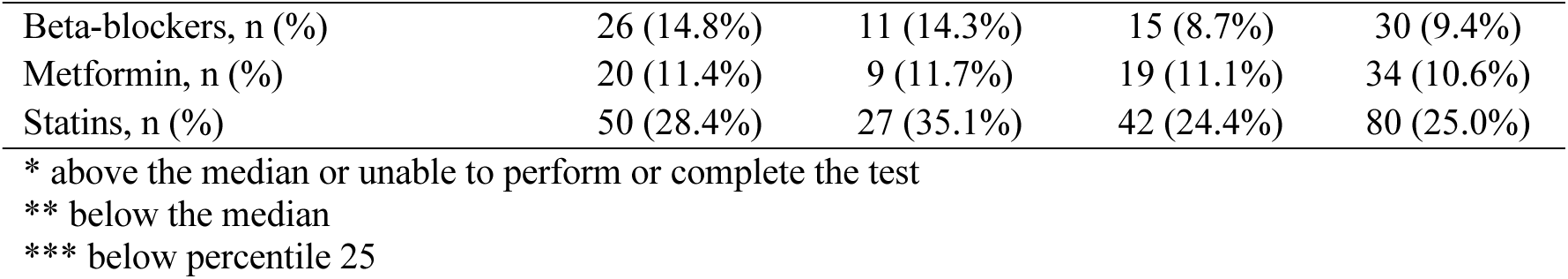
General characteristics of participants in the analysis of physical fitness related to exposure to PM_2.5_ in Medellin.

| Variable |  | Heart rate at rest<br>(inadequate*) | Muscle power<br>(inadequate*) | Grip strength<br>(inadequate*) | Total<br>Participants |
| --- | --- | --- | --- | --- | --- |
| n (%) |  | 176 (55.0) | 77 (24.0) | 172 (53.7) | 320 (100) |
| **Demographic** |  |  |  |  |  |
| Age (years), mean (SD) |  | 62 ± (9.0) | 61 ± (9.0) | 59 ± (9.0) | 60 ± (8.7) |
| Women, n (%) |  | 113 (75.6%) | 52 (67.5%) | 157 (91.3%) | 228 (71.2%) |
| Marital status married, n (%) |  | 81 (46.0%) | 36 (46.8%) | 73 (42.4%) | 150 (46.9%) |
| Sub-city unit, n (%) | 1 | 114 (64.8%) | 32 (41.6%) | 103 (59.9%) | 203 (63.4%) |
|  | 2 | 62 (35.2%) | 45 (58.4%) | 69 (40.1%) | 117 (36.6%) |
| Percentile of exposure to ambient PM<sub>2.5</sub>, n (%) | <P25 | 42 (23.9%) | 32 (41.6%) | 53 (30.9%) | 80 (25%) |
|  | P25 – 75 | 83 (47.2%) | 29 (37.7%) | 77 (44.8%) | 160 (50%) |
|  | >P75 | 51 (29%) | 16 (20.8%) | 42 (24.4%) | 80 (25%) |
| **Clinical** |  |  |  |  |  |
| Body-mass index (kg/m<sup>2</sup>), mean (SD) |  | 29.2 ± (5.4) | 29.7 ± (5.8) | 28.8 ± (5.6) | 28.2 ± (5.1) |
| Heart rate (beats/min), mean (SD) |  | 79 ± (11) | 76 ± (12) | 77 ± (10) | 77 ± (12) |
| Systolic blood pressure (mm Hg), mean (SD) |  | 123 ± (18) | 119 ± (15) | 119 ± (18) | 120 ± (17) |
| Diastolic blood pressure (mm Hg), mean (SD) |  | 77 ± (12) | 76 ± (11) | 72 ± (12) | 76 ± (12) |
| **Personal habits** |  |  |  |  |  |
| Current smokers, n (%) |  | 18 (10.2%) | 14 (18.2%) | 25 (14.5%) | 43 (13.4%) |
| Insufficient physical activity, n (%) |  | 116 (65.9%) | 55 (71.4%) | 115 (66.9%) | 198 (61.9%) |
| **Funcionales** |  |  |  |  |  |
| % Heart rate at rest, mean (SD) |  | 59.5 ± (14.8) | 44.1 ± (16.1) | 46.3 ± (17.3) | 45.8 ± (18.1) |
| Relative Muscle power (W·kg<sup>-1</sup>), mean (SD) |  | 2.6 ± (0.9) | 1.9 ± (0.6) | 2.4 ± (0.8) | 2.6 ± (1.0) |
| Grip strength (Kg), mean (SD) |  | 21.7 ± (7.4) | 21.4 ± (8.1) | 18.3 ± (4.9) | 23.0 ± (8.5) |
| **Morbidity** |  |  |  |  |  |
| Charlson comorbidity index, median (IQR) |  | 1.8 (0.9-2.9) | 1.5 (0.7-2.8) | 1.3 (0.5-2.6) | 1.5 (0.7-2.7) |
| Obesity, n (%) |  | 72 (40.9%) | 34 (44.2%) | 66 (38.4%) | 107 (33.4%) |
| High blood pressure, n (%) |  | 86 (48.9%) | 36 (46.8%) | 66 (38.4%) | 127 (39.7%) |
| Diabetes mellitus, n (%) |  | 24 (13.6%) | 10 (13.0%) | 20 (11.6%) | 43 (13.4%) |
| Acute myocardial infarction, n (%) |  | 11 (6.3%) | 3 (3.9%) | 6 (3.5%) | 13 (4.1%) |
| Heart failure, n (%) |  | 13 (7.4%) | 5 (6.5%) | 8 (4.7%) | 15 (4.7%) |
| Cerebrovascular disease, n (%) |  | 7 (4.0%) | 2 (2.6%) | 3 (1.7%) | 10 (3.1%) |
| Peripheral vascular disease, n (%) |  | 26 (14.8%) | 16 (20.8%) | 26 (15.1%) | 39 (12.2%) |
| Chronic obstructive pulmonary disease, n (%) |  | 11 (6.3%) | 6 (7.8%) | 10 (5.8%) | 16 (5.0%) |
| **Medications use** |  |  |  |  |  |
| Diuretics, n (%) |  | 29 (16.5%) | 16 (20.8%) | 24 (14.0%) | 42 (13.1%) |
| angiotensin-converting enzyme inhibitors, n (%) |  | 24 (13.6%) | 9 (11.7%) | 19 (11.1%) | 36 (11.3%) |
| Angiotensin 2 receptor blockers, n (%) |  | 58 (33.0%) | 24 (31.2%) | 45 (26.2%) | 83 (25.9%) |

|  |  |  |  |  |
| --- | --- | --- | --- | --- |
| Beta-blockers, n (%) | 26 (14.8%) | 11 (14.3%) | 15 (8.7%) | 30 (9.4%) |
| Metformin, n (%) | 20 (11.4%) | 9 (11.7%) | 19 (11.1%) | 34 (10.6%) |
| Statins, n (%) | 50 (28.4%) | 27 (35.1%) | 42 (24.4%) | 80 (25.0%) |

\* above the median or unable to perform or complete the test
\*\* below the median
\*\*\* below percentile 25

### Ambient PM_2.5_ exposure of the participant’s place of residence

Air pollution in Medellin has improved in the last years, with a 12 µg/m^3^ decrease in the annual average of PM_2.5_ between 2015 and 2023. For 2022, a LUR model was developed using the data of wind speed (WS), ambient temperature gradient (TG), and transportation route land use (TR)in the buffer 150m (PM2.5 = 25.97317 - 3.70696*WS - 70.50614*TG + 0.00014*TR) that explained 78% of the geographical variability of PM_2.5_, with a slight error in the predicted concentrations (root mean square error of 1.77). Based on this model, the annual concentrations of ambient PM_2.5_ were established in the residence address of the 320 study participants (Figure 2). Among those evaluated, a median of 19.61 µg/m^3^ was calculated, with values between 13.86 µg/m^3^ and 22.27 µg/m^3^ and IQR of 2.8.

**Figure 2.**
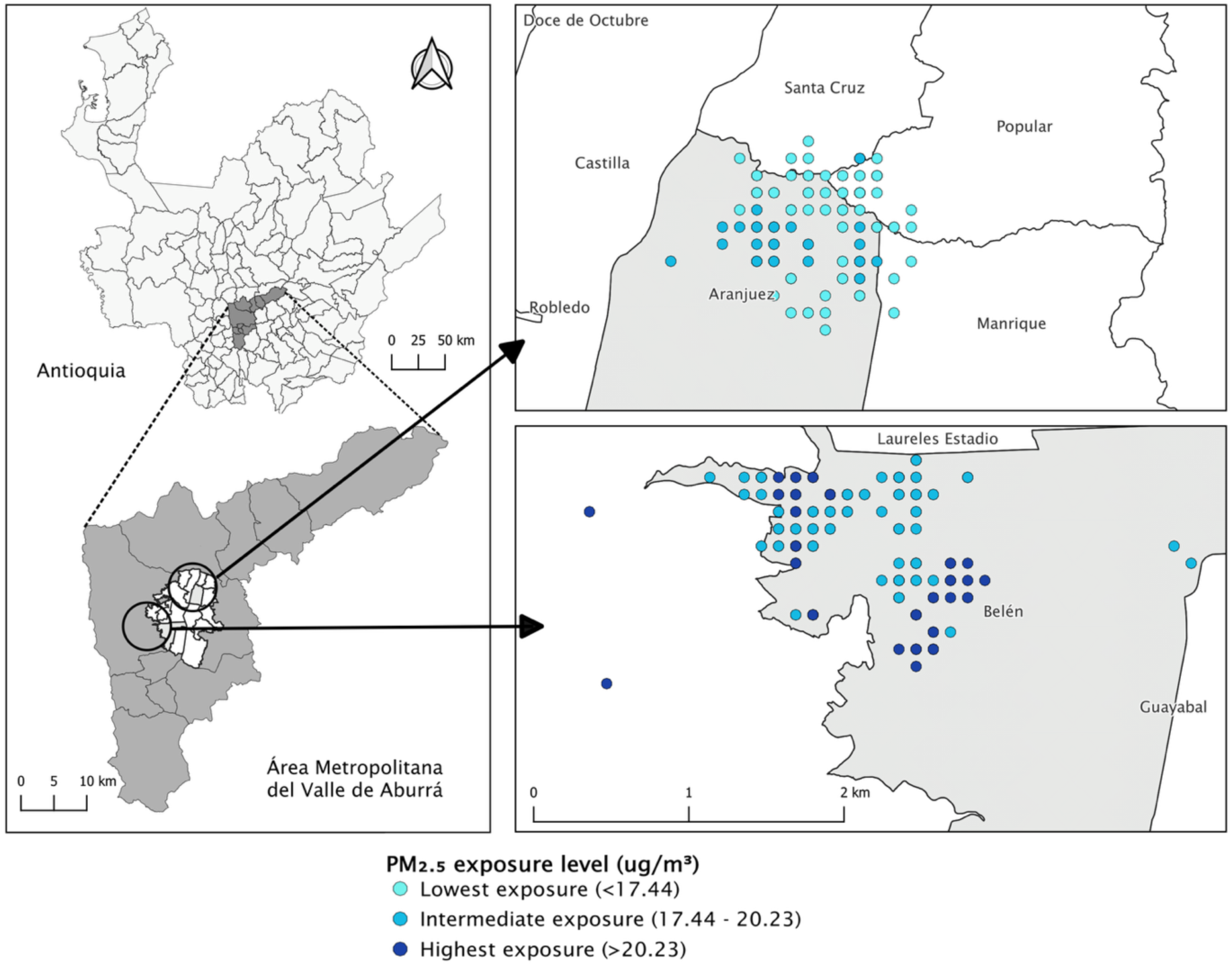
Location of participants in the study area and classification of their exposure according to percentiles: lowest exposure <P25, intermediate exposure between P25-P75, and highest exposure ≥P75

### Relationship between ambient PM_2.5_ exposure and PF

Higher use of the HR_r_ was observed in people with greater exposure to PM_2.5_ compared to the group with less exposure (ß coefficient = 7.11; 95% CI 1.11-13.12; p-value = 0.02). People with greater exposure had a better MF, expressed in power relative to weight (ß coefficient = 0.50; 95% CI 0.23-0.77; p-value < 0.01) and grip strength (ß coefficient = 3.59; 95% CI 1.63-5.56; p-value < 0.01). It should be noted that adjustments were made for age, BMI, sex, physical activity level, Charlson comorbidity index, socioeconomic stratum, and cigarette consumption (Table 2), which were consistent with the sensitivity analyses carried out (Table 3).

**Table 2.**
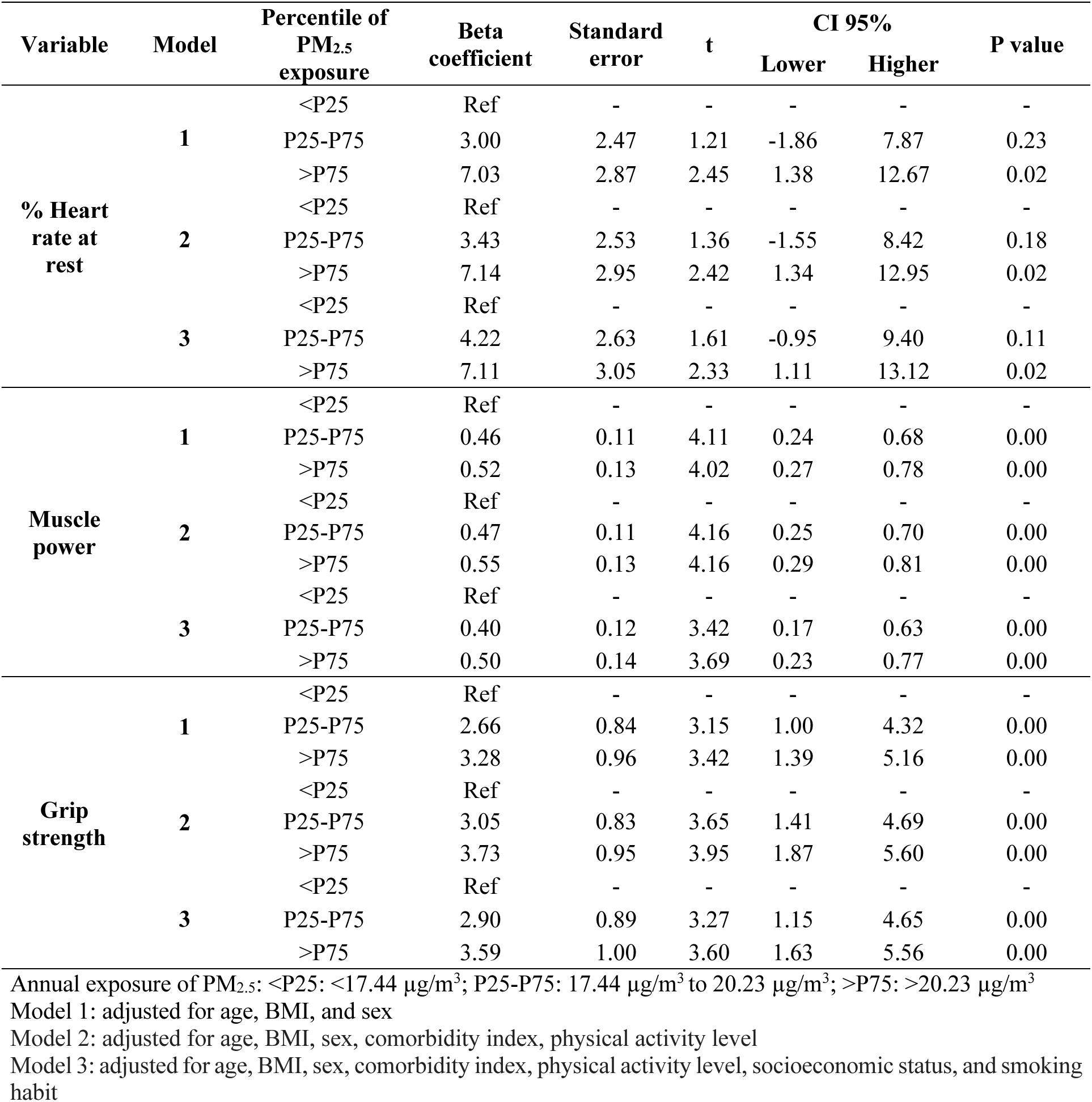
Multiple linear regression analysis between physical fitness indicators and the exposure to ambient PM_2.5_.

| Variable | Model | Percentile of PM<sub>2.5</sub> exposure | Beta coefficient | Standard error | t | CI 95% |  | P value |
| --- | --- | --- | --- | --- | --- | --- | --- | --- |
|  |  |  |  |  |  | Lower | Higher |  |
| % Heart rate at rest | 1 | <P25 | Ref | - | - | - | - | - |
|  |  | P25-P75 | 3.00 | 2.47 | 1.21 | -1.86 | 7.87 | 0.23 |
|  |  | >P75 | 7.03 | 2.87 | 2.45 | 1.38 | 12.67 | 0.02 |
|  | 2 | <P25 | Ref | - | - | - | - | - |
|  |  | P25-P75 | 3.43 | 2.53 | 1.36 | -1.55 | 8.42 | 0.18 |
|  |  | >P75 | 7.14 | 2.95 | 2.42 | 1.34 | 12.95 | 0.02 |
|  | 3 | <P25 | Ref | - | - | - | - | - |
|  |  | P25-P75 | 4.22 | 2.63 | 1.61 | -0.95 | 9.40 | 0.11 |
|  |  | >P75 | 7.11 | 3.05 | 2.33 | 1.11 | 13.12 | 0.02 |
| Muscle power | 1 | <P25 | Ref | - | - | - | - | - |
|  |  | P25-P75 | 0.46 | 0.11 | 4.11 | 0.24 | 0.68 | 0.00 |
|  |  | >P75 | 0.52 | 0.13 | 4.02 | 0.27 | 0.78 | 0.00 |
|  | 2 | <P25 | Ref | - | - | - | - | - |
|  |  | P25-P75 | 0.47 | 0.11 | 4.16 | 0.25 | 0.70 | 0.00 |
|  |  | >P75 | 0.55 | 0.13 | 4.16 | 0.29 | 0.81 | 0.00 |
|  | 3 | <P25 | Ref | - | - | - | - | - |
|  |  | P25-P75 | 0.40 | 0.12 | 3.42 | 0.17 | 0.63 | 0.00 |
|  |  | >P75 | 0.50 | 0.14 | 3.69 | 0.23 | 0.77 | 0.00 |
| Grip strength | 1 | <P25 | Ref | - | - | - | - | - |
|  |  | P25-P75 | 2.66 | 0.84 | 3.15 | 1.00 | 4.32 | 0.00 |
|  |  | >P75 | 3.28 | 0.96 | 3.42 | 1.39 | 5.16 | 0.00 |
|  | 2 | <P25 | Ref | - | - | - | - | - |
|  |  | P25-P75 | 3.05 | 0.83 | 3.65 | 1.41 | 4.69 | 0.00 |
|  |  | >P75 | 3.73 | 0.95 | 3.95 | 1.87 | 5.60 | 0.00 |
|  | 3 | <P25 | Ref | - | - | - | - | - |
|  |  | P25-P75 | 2.90 | 0.89 | 3.27 | 1.15 | 4.65 | 0.00 |
|  |  | >P75 | 3.59 | 1.00 | 3.60 | 1.63 | 5.56 | 0.00 |

Annual exposure of PM<sub>2.5</sub>: <P25: <17.44 µg/m<sup>3</sup>; P25-P75: 17.44 µg/m<sup>3</sup> to 20.23 µg/m<sup>3</sup>; >P75: >20.23 µg/m<sup>3</sup>
Model 1: adjusted for age, BMI, and sex
Model 2: adjusted for age, BMI, sex, comorbidity index, physical activity level
Model 3: adjusted for age, BMI, sex, comorbidity index, physical activity level, socioeconomic status, and smoking habit

**Table 3:** Sensitivity analysis of the relationship between exposure to ambient PM2.5 and fitness indicators (Binary logistic regression models)

| Variable | Model | Level of PM<sub>2.5</sub> exposure | Beta coefficient | Standard error | t | CI 95% |  | P value |
| --- | --- | --- | --- | --- | --- | --- | --- | --- |
| % Heart rate at rest | 1 | <P25 | Ref | - | - | - | - | - |
|  |  | P25-P75 | 1.13 | 0.33 | 0.44 | 0.64 | 2.02 | 0.65 |
|  |  | P75 | 1.77 | 0.61 | 1.66 | 0.90 | 3.48 | 0.09 |
|  | 2 | <P25 | Ref | - | - | - | - | - |
|  |  | P25-P75 | 1.23 | 0.37 | 0.68 | 0.68 | 2.22 | 1.49 |
|  |  | P75 | 1.81 | 0.64 | 1.68 | 0.94 | 3.63 | 0.09 |
|  | 3 | <P25 | Ref | - | - | - | - | - |
|  |  | P25-P75 | 1.32 | 0.43 | 0.84 | 0.69 | 2.50 | 0.40 |
|  |  | P75 | 1.70 | 0.65 | 1.41 | 0.81 | 3.59 | 0.15 |
| Muscle power | 1 | <P25 | Ref | - | - | - | - | - |
|  |  | P25-P75 | 0.30 | 0.09 | -3.71 | 0.16 | 0.57 | 0.00 |
|  |  | P75 | 0.33 | 0.12 | -2.84 | 0.16 | 0.71 | 0.00 |
|  | 2 | <P25 | Ref | - | - | - | - | - |
|  |  | P25-P75 | 0.29 | 0.09 | -3.62 | 0.12 | 0.57 | 0.00 |
|  |  | P75 | 0.31 | 0.12 | -2.90 | 0.14 | 0.68 | 0.00 |
|  | 3 | <P25 | Ref | - | - | - | - | - |
|  |  | P25-P75 | 0.29 | 0.09 | -3.62 | 0.12 | 0.57 | 0.00 |
|  |  | P75 | 0.31 | 0.12 | 2.90 | 0.14 | 0.68 | 0.00 |
| Grip strength | 1 | <P25 | Ref | - | - | - | - | - |
|  |  | P25-P75 | 0.38 | 0.13 | -2.63 | 0.29 | 0.78 | 0.00 |
|  |  | P75 | 0.44 | 0.17 | -2.04 | 0.20 | 0.96 | 0.04 |
|  | 2 | <P25 | Ref | - | - | - | - | - |
|  |  | P25-P75 | 0.31 | 0.11 | -2.04 | 0.14 | 0.66 | 0.00 |
|  |  | P75 | 0.34 | 0.14 | -2.54 | 0.15 | 0.78 | 0.01 |
|  | 3 | <P25 | Ref | - | - | - | - | - |
|  |  | P25-P75 | 0.29 | 0.12 | -2.97 | 0.13 | 0.66 | 0.00 |
|  |  | P75 | 0.31 | 0.14 | -2.7 | 0.13 | 0.76 | 0.01 |

Model 1: adjusted for age, BMI, and sex
Model 2: adjusted for age, BMI, sex, comorbidity index, physical activity level
Model 3: adjusted for age, BMI, sex, comorbidity index, physical activity level, socioeconomic status, and smoking habit

## DISCUSSION

Multiple publications relate exposure to PM_2.5_ and disease burden from non-communicable diseases [3], both for chronic respiratory [28] and cardiovascular outcomes. The risk of morbidity increases up to 20% for every 10 μg/m^3^ exposure to PM_2.5_ [29]. However, despite recognizing these associations, little has been done to analyze robust health indicators such as PF.

This study is one of the first in Latin America to analyze FP and long-term PM_2.5_ ambient air pollution exposure in the adult population. Although the air quality in Medellin has not reached the levels suggested by the WHO to guarantee good health (<5 µg/m^3^), it is on the right path to fulfilling provisional objectives and intermediate goals (intermediate goal No. 2: < 25 µg/m^3^) [2]. One of the study’s main findings was that people with greater exposure had a lower CRF and a better MF. As hypothesized, higher PM_2.5_ exposure resulted in a greater use of the %HR_r_, which supports other evidence that associates PM_2.5_ with chronic disease and mortality [7].

In the sample used in this study, it was identified that people with intermediate exposure used up to 4.22% more %HR_r_, while in the group with greater exposure, this increased to 7.14%. Several biological mechanisms generated by exposure to air pollutants can explain the decline in CRF, among which are pulmonary oxidative stress and systemic inflammation due to an increase in free radicals, an increase in blood pressure due to a decrease in nitric oxide, and a reduction in heart rate variability due to an increase in sympathetic tone in the autonomic nervous system [30].

Contrary to our hypothesis, we found that participants with greater pollutant exposure had a better MF. In contrast, recent studies have described how high exposure to pollutants has a negative impact on MF [31, 32]. For every 10 μg/m^3^ increase in ambient PM_2.5_ concentrations, a reduction in handgrip strength of 0.70 kg has been reported among people in developing countries [33]. This phenomenon is attributed to various mechanisms, including increased oxidative stress and inflammation directly affecting muscle mitochondrial function [32]. People exposed to high levels of environmental pollution could develop physiological adaptations.

The worst CRF in these scenarios forces individuals to compensate for this deficiency with muscular adaptations, increasing this capacity and the efficient use of oxygen [34]. Another mechanism that could help explain this finding is the “Fat but powerful” obesity paradox, given that studies have shown that overweight older adults have a greater relative power [35]. In our sample, there was a higher prevalence of obesity in the group with higher exposure to PM_2.5_ than in the group with lower exposure (37.5% vs. 26.2%). However, this finding is not sufficient to establish an association between the results. We must take into account the possibility of residual biases, such as the type of exercise performed by the participants. If the exercise were predominantly strength-focused, this capacity could be developed and give better test results. However, discrimination of the type of exercises was not asked of the participants.

This study stands out for being, to date, the first cross-sectional investigation nested in a population-based cohort (MEDELLIN Cohort) formed from multistage sampling. In Colombia, in the last two decades, research into the epidemiology of air pollution has focused on the relationship between environmental variables and health at the population level [12,13], with recent advances in the construction of small-area exposure models [36]. The methodology of the primary study helped control the selection bias and used tools that allowed for the analysis of geographical variation in PM_2.5_ concentrations and the assignment of individual exposure based on place of residence.

CRF and MF were determined through validated physical tests applied to each participant. Although the current standard method to evaluate CRF is to quantify oxygen consumption, the population-based clinical utility of indirect submaximal tests has been validated [37]. Considering the population of the study, this choice turned out to be safer and more practical.

One of the study’s limitations was that the final sample of participants was small compared to the total number of people admitted to the cohort, potentially incurring a selection bias that limited the extent to which the results could be generalized for other populations. The main limitation in recruiting participants was their lack of availability to attend the medical evaluation. While we cannot determine whether the evaluated participants had a better PF, we found similar characteristics between the sample and the selected cohort regarding age, place of residence, and sex. Another limitation was the determination of exposure based solely on place of residence. It is possible that participants work or spend time in areas other than where they reside, and these areas could have different levels of exposure. However, less than a quarter of participants were employed, suggesting that they spent more time at home.

Finally, as this was a cross-sectional study, we cannot establish a causal relationship between long-term exposure to PM_2.5_ and PF. The complexity of these interactions requires further analysis. This line of research should be continued to expand our understanding and develop effective strategies through physical activity and exercise to mitigate the adverse impact of particulate matter ambient air pollution exposure on human health.

## CONCLUSIONS

Our results show that people with greater PM_2.5_ ambient air pollution exposure have worse CRF and better MF. Although we recognize the limitations, these results support the need for new research to understand the impacts of air pollution on human health and develop effective interventions.

## ACKNOWLEDGEMENTS

The authors would like to thank the Health Department of Medellín and the environmental authority of the Aburrá Valley for their support during the data collection, and Santiago Mejía and Sara Grisales Vargas for their contributions to the development of the LUR model analysis. They would also like to thank the communities, organizations, and academic institutions that participated in the project and contributed with human, material, and location resources.

## DECLARATIONS

### Ethics approval

This research was approved by the Research Ethics Committee of the National Faculty of Public Health of the University of Antioquia in session 165 of May 23, 2017 (number: 21030002 – 00223 – 2019). All procedures followed the ethical standards of the Declaration of Helsinki and its 2013 adaptation.

### Consent to Participate

All participants agreed to participate voluntarily and signed an informed consent form previously approved by the National Faculty of Public Health ethics committee. This form allowed the follow-up of the cohort participants and authorized access to their medical records

## Authors contribution

*Duván Alexis Gómez-Castro*: Conceptualization, Methodology, Data collection-Investigation, Formal analysis, Writing - original draft preparation, Writing - review and editing. *Ana Liseth Herrera-Gómez*: Conceptualization, Methodology, Data collection- Investigation, Formal analysis, Writing - original draft preparation, Writing - review and editing. *Carolina Betancur-Figueroa*: Conceptualization, Methodology, Data collection- Investigation, Writing - original draft preparation, Writing - review and editing. *Juan Gabriel Piñeros-Jiménez*: Conceptualization, Methodology, Writing - review and editing, Funding acquisition, Resources, Supervision. *Jaime Alberto Gallo-Villegas*: Conceptualization, Methodology, Formal analysis, Writing - review and editing, Supervision

## Data availability

The data generated in this research is stored in the Research Data Repository Zenodo. Dataset Exposure to PM2.5 ambient air pollution and fitness [Data set]. Zenodo. https://doi.org/10.5281/zenodo.14783011 (Gómez-Castro et al. 2025).

## Consent for Publication

Not applicable.

## Code Availability

Not applicable

## Trial registration

Not applicable

